# Bibliometric analysis of COVID-19 literature

**DOI:** 10.1101/2020.07.15.20154989

**Authors:** Md. Sayeed Al-Zaman

## Abstract

This study analyzes *N*=16384 COVID-19-related literature published between December 2019 to June 2020. The data were extracted from the Web of Science database using four keywords: “COVID-19”, “Coronavirus”, “2019-nCoV”, and “SARS-CoV-2”. The analysis found that almost all but a tiny number of the papers are published in 2020 (95.16%). Of the 15 types of publications, *article* (40.015%) is on the top of the list. All publications are in 19 different languages where English (95.313%) is the dominant one. A total of 159 countries produce COVID-19-related researches, and the USA (25.433%) is in the leading position. According to the findings, Wang Y (*n*=94) is the top author, and the British Medical Journal (BMJ) (*n*=488) is the top source. Also, the University of London (*n*=488) is the leading organization with the highest number of papers, and medicine-related papers (*n*=2259) are the highest in numbers. Apart from these novel findings, this study is perhaps the largest COVID-19-related bibliometric analysis to date.

## Introduction

The first COVID-19 case was identified on 17 November 2019 in Wuhan, the capital of the Hubei province of China (Darsono et al., 2020). The virus reached at least 25 countries as of 6 February 2020 and became global soon after (Wu et al., 2020). The World Health Organization (WHO) immediately named it “COVID-19” and declared the situation as a *pandemic*. With time, the virus engulfed most of the countries, and the infection fatality rate (IFR) surged. From 1 December 2019 to 14 July 2020, 13.25 million people got infected in 215 countries: 0.58 million people died, 7.72 million people recovered, and 4.96 million cases are still active (Worldometer, 2020). In many countries like Italy, the USA, the UK, Spain, and China, the spread experiences a decrease while in many countries like Bangladesh and India, the infection rate is still surging. Amid such a situation, the more deadly “Second Wave” of the pandemic is expected to come at the end of 2020 (Roberts, 2020). Thanks to the pandemic, researchers from various fields are flooding academia with thousands of publications: some are medicine-related while some are healthcare- and virology-related. Having many positive contributions, this trend has some drawbacks since many published papers are found as unsubstantial. In a recent analysis, for example, Dr. Elisabeth Bik, an image-analyst and a former Stanford researcher, revealed how many researchers are reusing “identical set of images” in their published papers (Xiao, 2020). Apart from such uninvited issues, understanding the ongoing COVID-19-related research trend is essential, and a systematic bibliometric analysis of the relevant published literature may able to provide some insights in this respect. Bibliometric analysis is the systematic statistical analysis of the published scholarly works, such as of articles, of books, of book chapters, and of reviews, to measure their impacts in the scientific arena (Iftikhar et al., 2019). Meanwhile, several COVID-19-related bibliometric papers added some relevant findings to the international scholarships. While some researches took only the recent publications into account (Golinelli et al., 2020; Lou et al., 2020; Zhai et al., 2020), some researches analyzed publications from a relatively longer span (Tao et al., 2020; Zhou & Chen, 2020). Identifying some knowledge-gaps in the previous literature, this bibliometric analysis presents some novel findings. The following discussion is divided into four consecutive sections. A few key studies are discussed in the literature review section. The processes of data collection and analysis are included in the method section. The result section presents the major and minor results from the analysis. The remarkable findings are presented and compared with the previous findings in the discussion section.

## Literature Review

From December 2019 to June 2020, COVID-19-related literature surges in an unprecedented manner. Following this surge of scholarly outputs, it becomes important to understand the research trend. To provide so, at least 15 relevant bibliometric analyses are published to date. These studies share some common structures, analytical aspects, and outputs. For example, most of the study used one or more similar but specific keywords to search PubMed, Scopus, and Web of Science databases for relevant literature within a specific time. A few key studies might represent the overall picture of the ongoing research trend around the world.

Using keyword searches, Kambhampati et al. (2020) extracted the PubMed data for 6831 papers. They found 6415 papers published in English; 1802 publications were human studies-related. They also found that the COVID-19-related papers grew exponentially during the period. A total of 1430 journals published papers related to the pandemic, and the British Medicine Journal (BMJ) published the highest 252 papers, followed by the Journal of Medical Virology (186 papers). Instead of producing countries, they searched for the names of the countries mentioned in the titles. They found that China was mentioned in 438 papers, followed by Italy in 127 papers. In their analysis, they found *review articles* were the most common document type with 202 papers. However, this study has a few limitations. For example, it only mentioned about the highest producing country, language, type, and source, but did not extend the results and explanation, such as the number of countries, types, and languages of the publications. Moreover, the paper did not mention about its data collection period that makes it difficult to understand the context.

Focusing on information and knowledge sharing among the international COVID-19 researchers, in a separate study, Darsono et al. (2020) collected bibliometric data of 1475 publications from the Scopus database. All papers were published between December 2019 to March 2020. Total 801 (45.3%) and 674 (45.7%) papers were published in 2019 and 2020, respectively. The study found 11 types of publications. Of them, *articles* had the highest 66.1% papers, followed by reviews (11.3%) and notes (7.6%). According to this study, Viruses was the leading journal with 74 published papers, followed by Lancet (50 papers) and the Journal of Virology (39 papers). It also found that Z. A. Memish (17 papers), E. Mahase (15 papers), and J. A. Al-Tawfiq (14 papers) were the top three authors in terms of published papers. China as a country produced the highest 386 papers and the University of Hong Kong as an organization produced the highest 44 papers, according to the study. Evaluating the findings, it seems the study focused more on the Islamic world.

In another similar study, Dehghanbanadaki et al. (2020) using keyword search extracted data of 923 COVID-19 documents indexed in the Scopus. The data collection period was chosen from 1 December 2019 to 1 April 2020. The data included “document type, open accessibility status, citation counting, H-index, top-cited documents, the most productive countries, institutions and journals, international collaboration, the most frequent terms and keywords, journal bibliographic coupling and co-citations” (Dehghanbanadaki et al., 2020, p. 354). In the country list, China ranked the first with 348 documents, followed by the USA with 160 documents. The Lancet and BMJ Research Ed published the highest number of publications (each with 74 documents). Both the University of Hong Kong and Huazhong University of Science and Technology ranked the first with 30 publications.

Tao et al. (2020) conducted a bibliometric analysis with a relatively larger dataset than the previously discussed studies. Unlike other studies, however, this study dealt with a longer period as well, i.e., 20 years. The researchers extracted 9760 publications’ data from 2000 to 2020. The source of data was the Web of Science database. The Journal of Virology was on the top in this field over the period with 885 publications. The USA was a leading country with 959 publications, followed by China with 469 publications. The University of Hong Kong was the highest-producing institution with 411 publications. Furthermore, Yuen KY and Peiris JSM were the two most productive researchers with 200 and 134 publications, respectively.

Lou et al. (2020) in their study searched the PubMed database with the keyword “COVID-19” and extracted the data of 183 publications published from 14 January to 29 February 2020. The data included title, corresponding author, language, publication time, publication type, and research focus. The result showed that the authors were from 20 different countries. Of them, 78 (42.6%) were from hospitals, 64 (35%) from universities, and 39 (21.3%) from research institutions. Eighty different journals published all the papers. The Journal of Medical Virology published the highest 25 papers. Most of the publications were articles (32.8%) while 15.8% were review and 10.9% were short communications. English was the dominant language, followed by Chinese. The result further showed the disciplines of the publications: 37.2% were in epidemiology, 26.8% in virology, and 14.2% in clinical features.

Some other relevant bibliometric studies (Chahrour et al., 2020; Chen et al., 2020; DE Felice & Polimeni, 2020; Golinelli et al., 2020; Hamidah et al., 2020; Hossain, 2020; Zhai et al., 2020; Zhou & Chen, 2020) analyzed the similar indices, such as authors, countries, languages, citations, institutions, sources, and types. These studies are affected by at least two problems. First, most of the studies analyzed literature from a relatively smaller span, and if not so, then with smaller samples. Therefore, these studies hardly present a broader and representative picture of the COVID-19 research trend. Second, as most of the researches dealt with smaller samples and span, they were likely to produce contradictory results. For example, Kambhampati et al. (2020) found BMJ as the highest-producing journal while Darsono et al. (2020) found the Lancet as the highest-producing journal. However, different research aims of the papers also could be responsible for such different results.

## Materials and Method

The bibliographic data for this research were collected from the Web of Science database, which is a popular database for data extraction for bibliometric analysis (e.g., the study of Tao et al. (2020)). The data processing is divided into three phases. *First*, the relevant literature was searched with four selected keywords and the Boolean “OR”: “COVID-19”, “Coronavirus”, “2019-nCoV”, and “SARS-CoV-2”. The search included the title, abstract, and author’s keywords. The keywords were determined based on the previous studies. For example, Darsono et al. (2020) used “Coronavirus” and “COVID-19” to search the literature while Kambhampati et al. (2020) used “Sar-Cov-2” and “COVID-19”. Among all the previous studies, however, Dehghanbanadaki et al. (2020) used the highest 23 keywords to search the Scopus database for available COVID-19-related literature. Note that the data collection period of the present study was longer than the previous studies. From December 2019 to June 2020, *N*=16384 scholarly publications appeared in different sources. In December 2019, 793 papers were published that was 4.84% of the total share (Table 01). However, the publication number surged next year. From January to June 2020, 15591 papers were published that was 95.16% of the total share. *Second*, the data were downloaded from the database in the *.txt* file format. It was transformed, restructured, and imported in IBM SPSS Statistics 25 for the final analysis. *Third*, the data processing had two tiers: general analysis and top percentile analysis. The two indices of the general analysis were the types and languages of the published papers. The five indices of the top percentile analysis were the top 10 authors, sources, countries, organizations, and disciplines of the published papers. In percentile indices, total 55352 authors, 2964 sources, 159 countries, 12805 organizations, and 221 disciplines were found for 16384 published papers.

**Table 01:**
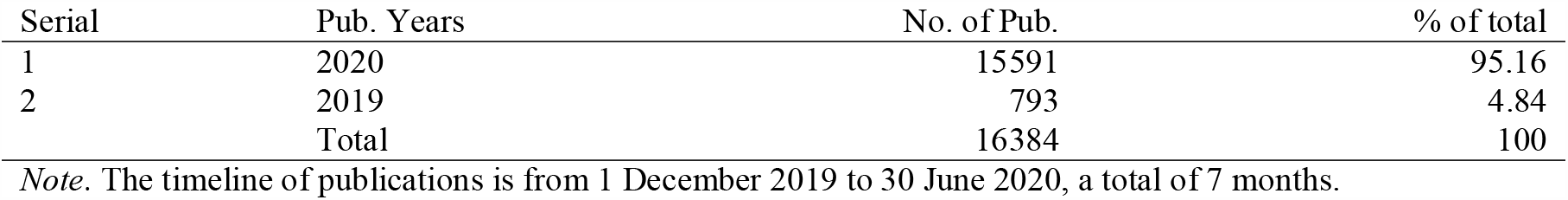
Overview of the publications.

| Serial | Pub. Years | No. of Pub. | % of total |
| --- | --- | --- | --- |
| 1 | 2020 | 15591 | 95.16 |
| 2 | 2019 | 793 | 4.84 |
|  | Total | 16384 | 100 |

*Note.* The timeline of publications is from 1 December 2019 to 30 June 2020, a total of 7 months.

## Results

A total of 15 document types are found (Table 02). Of them, *article* has the highest share (*n*=6556; 40.015%), followed by editorial material (*n*=4138; 25.256%). It is important to mention that many papers from a few categories often overlap which may increase the sum of the papers. For example, an *article* or a *review* can also be an *early access* paper. Papers are published in 19 different languages (Table 03). Of them, papers in English are unevenly higher than the others that is *n*=15616 (95.313%). German (*n*=203; 1.239%) and Spanish (*n*=196; 1.196%) languages occupy the second and third positions on the list, respectively. Catalan, Croatian, Icelandic, and Indonesian languages are on the bottom of the list having only *n*=1 (0.006%) paper.

**Table 02:** Types of the publications.

| Rank | Document Types | No. of Pub. | % of total |
| --- | --- | --- | --- |
| 1 | Article | 6556 | 40.015 |
| 2 | Editorial Material | 4138 | 25.256 |
| 3 | Early Access | 3878 | 23.669 |
| 4 | Letter | 3398 | 20.74 |
| 5 | Review | 1502 | 9.167 |
| 6 | News Item | 631 | 3.851 |
| 7 | Correction | 130 | 0.793 |
| 8 | Meeting Abstract | 20 | 0.122 |
| 9 | Data Paper | 13 | 0.079 |
| 10 | Book Chapter | 7 | 0.043 |
| 11 | Book Review | 4 | 0.024 |
| 12 | Biographical Item | 3 | 0.018 |
| 13 | Proceedings Paper | 3 | 0.018 |
| 14 | Dance Performance Review | 1 | 0.006 |
| 15 | Reprint | 1 | 0.006 |

*Note.* The list includes all the types of the published papers instead of selected few.

**Table 03:** Languages of the publications.

| Rank | Languages | No. of Pub. | % of total |
| --- | --- | --- | --- |
| 1 | English | 15616 | 95.313 |
| 2 | German | 203 | 1.239 |
| 3 | Spanish | 196 | 1.196 |
| 4 | Italian | 105 | 0.641 |
| 5 | French | 90 | 0.549 |
| 6 | Portuguese | 39 | 0.238 |
| 7 | Norwegian | 32 | 0.195 |
| 8 | Hungarian | 31 | 0.189 |
| 9 | Russian | 24 | 0.146 |
| 10 | Turkish | 24 | 0.146 |
| 11 | Korean | 5 | 0.031 |
| 12 | Polish | 5 | 0.031 |
| 13 | Chinese | 4 | 0.024 |
| 14 | Czech | 3 | 0.018 |
| 15 | Dutch | 3 | 0.018 |
| 16 | Catalan | 1 | 0.006 |
| 17 | Croatian | 1 | 0.006 |
| 18 | Icelandic | 1 | 0.006 |
| 19 | Indonesian | 1 | 0.006 |

*Note.* The list includes all the languages of the published papers instead of selected few.

The top 10 authors are 0.02% of the total authors, and they produce 5.663% (*n*=928) of the total outputs (Table 04). Of them, a remarkable number of authors are anonymous (*n*=282; 1.721%). Wang Y (*n*=94) produces the highest number of papers, followed by Zhang Y (*n*=88) and Li Y (*n*=77). The top 10 sources of publications constitute 0.34% of the total sources. They produce *n*=1974 papers that is 12.049% of the total publications (Table 05). Of them, the BMJ (*n*=488) publishes the highest number of papers, followed by the Journal of Medical Virology (*n*=303) and Journal of Infection (*n*=261). The top 10 countries constitute 6.29% of the total countries. Unlike the top 10 authors and sources, the top 10 countries produce the bulk share of the publications, i.e., *n*=14633 papers and 89.312% of the total outputs (Table 06). In the list, the USA secures the leading position with *n*=4167 published papers, followed by China (*n*=2979) and Italy (*n*=1921). It is to mention that a few previous studies explored the wide collaboration network among countries. For example, Dehghanbanadaki et al. (2020) identifies a total of 241 collaborations between China and the USA, which is the strongest of all. For such reasons, many publications may share two or more countries at the same time.

**Table 04:**
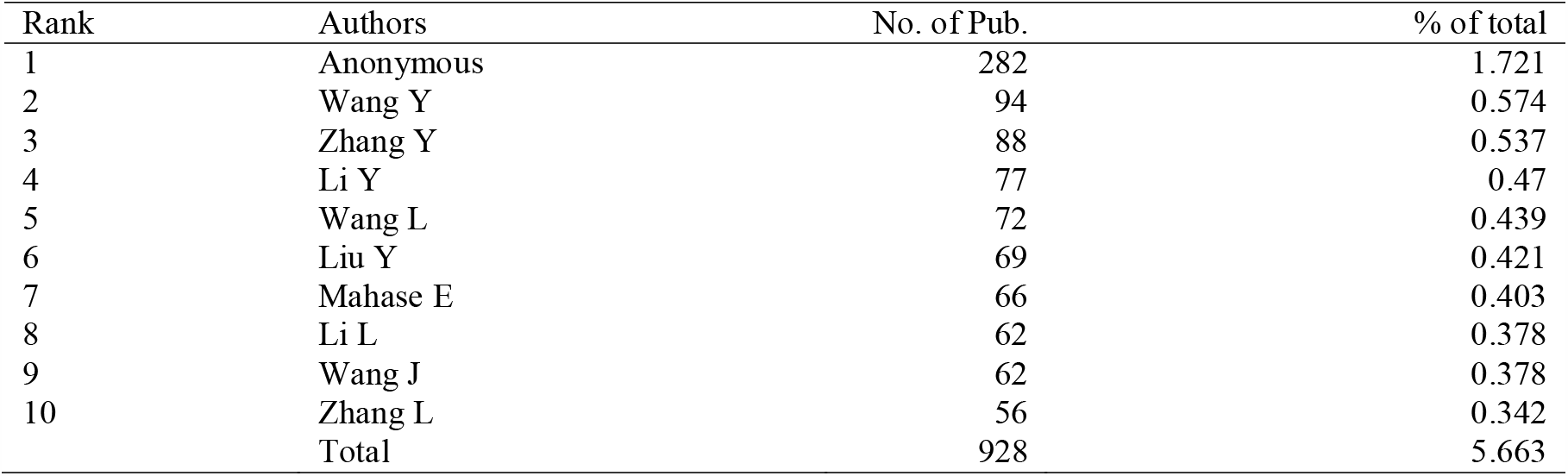
Top 10 authors.

**Table 05:** Top 10 sources.

| Rank | Source Titles | No. of Pub. | % of total |
| --- | --- | --- | --- |
| 1 | British Medical Journal (BMJ) | 488 | 2.979 |
| 2 | Journal of Medical Virology | 303 | 1.849 |
| 3 | Journal of Infection | 261 | 1.593 |
| 4 | Lancet | 191 | 1.166 |
| 5 | Cureus | 154 | 0.94 |
| 6 | Nature | 136 | 0.83 |
| 7 | Critical Care | 120 | 0.732 |
| 8 | Jama Journal of The American Medical Association | 113 | 0.69 |
| 9 | New England Journal of Medicine | 113 | 0.69 |
| 10 | Head and Neck | 95 | 0.58 |
|  | Total | 1974 | 12.049 |

**Table 06:** Top 10 countries.

| Rank | Country | No. of Pub. | % of total |
| --- | --- | --- | --- |
| 1 | USA | 4167 | 25.433 |
| 2 | China | 2979 | 18.182 |
| 3 | Italy | 1921 | 11.725 |
| 4 | England | 1575 | 9.613 |
| 5 | Germany | 745 | 4.547 |
| 6 | India | 738 | 4.504 |
| 7 | Canada | 730 | 4.456 |
| 8 | France | 662 | 4.041 |
| 9 | Australia | 620 | 3.784 |
| 10 | Spain | 496 | 3.027 |
|  | Total | 14633 | 89.312 |

The top 10 organizations are 0.08% of the total organizations. They cumulatively produce *n*=2895 papers, that is, 17.67% of the total outputs (Table 07). Of them, the University of London (*n*=488) has the highest outputs; Harvard University (*n*=403) and the University of California, Berkeley (*n*=352) are on the second and third position on the list, respectively. The top 10 popular disciplines are 4.53% of the total disciplines. They produce *n*=8814 papers that is 53.797% of the total outputs (Table 08). Of them, medicine-related papers (*n*=2259) are the highest in numbers, followed by public environment-related (*n*=1203) and infectious disease-related papers (*n*=1146).

**Table 07:** Top 10 organizations.

| Rank | Organizations | No. of Pub. | % of total |
| --- | --- | --- | --- |
| 1 | University of London | 488 | 2.979 |
| 2 | Harvard University | 403 | 2.46 |
| 3 | University of California System | 352 | 2.148 |
| 4 | Huazhong University of Science Technology | 339 | 2.069 |
| 5 | Harvard Medical School | 238 | 1.453 |
| 6 | Wuhan University | 220 | 1.343 |
| 7 | University College London | 218 | 1.331 |
| 8 | Chinese Academy of Sciences | 217 | 1.324 |
| 9 | Inserm | 216 | 1.318 |
| 10 | University of Toronto | 204 | 1.245 |
|  | Total | 2895 | 17.67 |

**Table 08:** Top 10 disciplines.

| Rank | Disciplines | No. of Pub. | % of total |
| --- | --- | --- | --- |
| 1 | Medicine General Internal | 2259 | 13.788 |
| 2 | Public Environmental Occupational Health | 1203 | 7.343 |
| 3 | Infectious Diseases | 1146 | 6.995 |
| 4 | Surgery | 827 | 5.048 |
| 5 | Virology | 733 | 4.474 |
| 6 | Immunology | 612 | 3.735 |
| 7 | Cardiac Cardiovascular Systems | 531 | 3.241 |
| 8 | Oncology | 525 | 3.204 |
| 9 | Medicine Research Experimental | 497 | 3.033 |
| 10 | Pharmacology Pharmacy | 481 | 2.936 |
|  | Total | 8814 | 53.797 |

## Discussion and Conclusion

This bibliometric study analyzed *N*=16384 publications’ data related to COVID-19 that were extracted from the Web of Science database. The papers were published between December 2019 to June 2020. It may be the largest sample of COVID-19-related bibliometric analysis to date. The analysis presents some novel findings. *First*, the data contain 15 types of publications. Of them, *article* is on the top with the highest publications, followed by editorial materials. Some previous studies also found the same but followed by either review and notes (Darsono et al., 2020) or review and short commentaries (Lou et al., 2020). However, some studies found reviews as the most popular type of publications (Kambhampati et al., 2020). *Second*, all papers are published in 19 different languages and most of them are in English, followed by German and Spanish. In their studies, Kambhampati et al., (2020) and Lou et al. (2020) also found that English is the dominant language of the published papers. However, in the second study, the researchers found Chinese as the second language on the list after English. *Third*, the 10 leading countries produce nine-tenth of the total publications. The USA is the leading country with the highest publications, followed by China. It supports the finding of Tao et al. (2020). (It is important to note that Tao’s study incorporates the data of 20 years, as mentioned earlier.) However, Darsono et al. (2020) and Dehghanbanadaki et al. (2020) produced a contradictory result showing China as the leading country. *Fourth*, the 10 leading authors produce only a small number of papers. However, although surprising, a remarkable number of authors are identified. Wang Y is found as the leading researcher, whereas Darsono et al. (2020) and Tao et al. (2020) found Z. A. Memish and Yuen KY as the leading authors, respectively. *Fifth*, the present study finds BMJ as the leading source of COVID-19-related researches, followed by the Journal of Medical Virology that supports the finding of Kambhampati et al. (2020) and contradicts the results of Darsono et al. (2020) and Lou et al. (2020). *Sixth*, the University of London produces the highest number of papers, followed by Harvard University. In contrast, Darsono et al. (2020), Dehghanbanadaki et al. (2020), and Tao et al. (2020) found the University of Hong Kong as the top-producing institute. The leading institutes also hint about the three contemporary research hotspots based on the UK, the USA, and China. *Seventh*, medicine-related publications are the highest in the list of disciplines, followed by environmental health-and infectious disease-related publications. It suggests that researchers more emphasize on COVID-19 medicine researches. While this study seeks a more specific field of research, Lou’s et al. (2020) result shows a few broader disciplines where epidemiology is on the leading position. Overall, this bibliometric analysis analyzing a large dataset produces some novel results that would guide further research in this area. It may further assist our understanding of the current trend of COVID-19-related researches from various angels and show some lights for the future researches. This study also hypothesizes that the production rate of the countries and their casualties inflicted by the pandemic are somewhat positively correlated in any extent. For example, the USA is the most affected country so is the leading producer of researches. Similarly, the UK and Italy are on the third and fifth positions on the casualty list (Worldometer, 2020) and fifth and third positions on the most-produced country list (see Table 06), respectively. Further studies might explore these areas as well.

## Data Availability

N/A

